# Application of the IASP grading system for ‘nociplastic pain’ in chronic pain conditions: A field study

**DOI:** 10.1101/2022.12.06.22283114

**Authors:** Hannah Schmidt, Armin Drusko, Malika Renz, Lea Schlömp, Heike Tost, Jonas Tesarz, Sigrid Schuh-Hofer, Andreas Meyer-Lindenberg, Rolf-Detlef Treede

## Abstract

The concept ‘nociplastic pain’ has been developed for patients in whom clinical and psychophysical findings suggest a predominant central sensitization type of pain that is not fully explained as nociceptive or neuropathic. Here we tested, how well the recently published grading system differentiates between chronic primary pain or chronic secondary pain conditions. We recruited patients with Fibromyalgia (FMS, 41), Complex Regional Pain Syndrome (CRPS, 11), Osteoarthritis (OA, 21) or Peripheral Nerve Injury (PNI, 8). We used clinical history, pain drawings, Quantitative Sensory Testing (QST) and questionnaires to classify patients’ pains as possibly or probably ‘nociplastic’ in nature.

All FMS and CRPS patients exhibited widespread or regional pain that was not explainable by nociceptive nor neuropathic mechanisms. Widespread pain in 12 OA patients was fully explained as nociceptive and regional pain in 4 PNI patients as neuropathic in all but one in each group. QST provided evidence for hypersensitivity in 9/11 CRPS patients but only 27/41 FMS patients (possible ‘nociplastic pain’). 82% of the CRPS patients but only 54% of FMS patients reported a history of hypersensitivity and mental comorbidities (probable ‘nociplastic pain’). We suggest that clinical examination of hypersensitivity should be done in more than one region and that adding a high tender point count as evidence for hypersensitivity phenomena may be useful. Further we suggest to switch the sequence of steps so that self-reported hypersensitivity and comorbidities come before clinical examination of hypersensitivity; Since the ‘nociplastic pain’ concept calls for brainstem and cortical plasticity we discuss in detail potential measurement strategies.

## 1. Introduction

The current edition of the International Statistical Classification of Diseases and Related Health Problems (ICD-11; WHO 2018 and 2022) distinguishes between chronic primary pain conditions (“chronic pain as a disease”) including Fibromyalgia Syndrome (FMS), Complex Regional Pain Syndrome (CRPS) and migraine headaches on one hand, and chronic secondary pain conditions on the other hand (“chronic pain as a symptom”): cancer-related pain, postsurgical or posttraumatic pain, neuropathic pain, secondary orofacial pain or headache, secondary visceral pain, secondary musculoskeletal pain. Altered nociceptive signal processing in the central nervous system (CNS) has been considered to play a role in the pathophysiology of all chronic primary pain conditions (Arendt-Nielsen & Graven-Nielsen, 2003; Fitzcharles et al., 2021), but they all fail to meet the current stringent neurological definition of neuropathic pain (Finnerup et al., 2016) that is also included in ICD-11.

Therefore, a task force of the International Association for the Study of Pain (IASP) proposed to add a third mechanistic descriptor called ‘nociplastic pain’ to the existing categories of nociceptive and neuropathic pain (Kosek et al., 2016). The rationale was to acknowledge mounting evidence that sensitization of spinal neurons, aberrant endogenous pain modulation, and functional as well as structural cortical network alterations seem to favor and maintain a chronic pain state in absence of tissue injury or somatosensory system lesions (Kosek et al., 2021; Nijs et al., 2021). This classification builds upon earlier proposals to label a „central sensitization type pain” (Nijs et al., 2014; Nijs et al., 2021).

Recently a grading system was published to identify chronic nociplastic pain in musculoskeletal conditions (Kosek et al., 2021). This grading system uses a classification tree based on evidence for widespread or regional rather than discrete pain distribution, hypersensitivity to external stimuli, and psychological symptoms named as comorbidities. Nociceptive or neuropathic pain conditions that would fully explain such patterns are exclusion criteria. The concepts of “central sensitization type pain” and ‘nociplastic pain’ have been criticized on theoretical grounds as being too broad to be useful (Cohen, 2022; Magerl, 2022) nociceptive signal processing in the CNS may be altered on at least three different levels (spinal, brainstem, thalamo-cortical; Treede et al., 2022) with different clinical manifestations, and at least spinal synaptic plasticity is a normal physiological consequence of any type of injury (“phase 2 pain” in Cervero & Laird, 1991). But the grading system is a major step forward because its classification results as possible or probable ‘nociplastic pain’ are testable empirically.

We tested the applicability of the proposed grading in a single-center study that is part of a larger collaborative research center (SFB 1158). We contrasted patients with one of two chronic primary pain conditions (FMS or CRPS) versus patients with one of two chronic secondary pain conditions: osteoarthritis (OA) or peripheral nerve injury (PNI). Our operationalization of the decision tree included medical history, questionnaires, pain drawings and quantitative sensory testing. We tested the hypothesis that the decision tree would assign patients with FMS or CRPS as probable nociplastic, and patients with OA or PNI as not nociplastic in a categorical classification.

## 2. Methods

The SUPER study had been ongoing before publication of the decision tree for musculoskeletal nociplastic pain (Kosek et al. 2021), but we noticed that in this framework we had recorded all variables needed to apply the proposed grading criteria to a cohort of chronic primary and secondary pain participants. In the following sections we give an overview about the SUPER study in general and the measures we used for the operationalization of the grading system in the here presented field test.

### 2.1 Participants

The participants for the two main groups chronic primary (CP1) vs chronic secondary pain (CP2) including two sub-diagnoses each (FMS and CRPS on the one hand and OA and PNI on the other hand) were recruited from the Rhine-Neckar-region in Germany between January 2020 and June 2022 via the ongoing SUPER study of SFB1158 (clinicaltrials.gov No: DRKS00029050). Besides these groups the SUPER framework also includes healthy controls and subjects suffering from Major Depression. FMS patients were mostly referred from the Department of Psychosomatic Medicine, Heidelberg, patients with CRPS, PNI or OA from the Department of Hand Surgery, BG clinic Ludwigshafen (Prof. Dr. Leila Harhaus). In addition, patients were recruited via patient advocacy groups and social media advertisements. The total sample size of the analysis reported here is 81 participants (see Tab 1). General in-and exclusion criteria for the chronic pain groups were at least 18 years of age and ability to give informed consent. Further, the pain must have been reported for at least 3 months before inclusion and the average pain should be of at least moderate severity (numeric rating scale (NRS) ≥ 4/10). For the CP1 groups in- and exclusion criteria were a confirmed diagnosis of FMS or CRPS based on documentation in patient records that followed established diagnostic criteria (Andreas Goebel et al., 2017; Finnerup et al., 2016; F. Wolfe et al., 1990; Frederick Wolfe et al., 2010) and freedom from specific rheumatological or systemic inflammatory or other major medical conditions. For inclusion in the CP2 group participants needed to present with a diagnosis of either a peripheral nerve injury (PNI; confirmed by a physician based on examination, nerve conduction studies or surgical evidence, following diagnostic steps proposed in (Finnerup et al., 2016) or a specific pathology of the spine (Osteo arthritis (OA) confirmed by a physician based on X-ray or MRI). All participants provided written informed consent before participation and were reimbursed with 200 Euro plus travel expenses. The study was approved by the local ethics committee (AZ 2018-663N-MA).

### 2.2 Study protocol

SUPER is an ongoing case-control study with two on-site days (∼ 4.5 hours on day 1 and day 8, respectively) at the Central Institute of Mental Health (CIMH), Mannheim, Germany, and 7 days of smartphone-based ecological momentary assessment (EMA) in-between. Interested participants were first screened by telephone for in- and exclusion criteria. On-site testing included a consent session, a structured clinical interview (Mini-DIPS; Margraf & Cwik, 2017) with a trained psychologist, a separate trauma interview, venipuncture as well as two neuroimaging (fMRI + Neurofeedback) and two neurophysiological sessions (one per on-site visit, respectively). The latter included Quantitative sensory testing (QST; Margraf & Cwik, 2017) conditioned pain modulation (CPM) and a high-frequency stimulation paradigm (HFS) as well as a sensory conditioning and a Jumping-to-conclusions-task. Moreover, in the neurophysiological sessions, ongoing pain characteristics were examined by pain drawings as well as questionnaires assessing neuropathic (nociplastic) components: DN4 (Bouhassira et al., 2005) and painDETECT (Freynhagen et al., 2016).

Additionally, the participants filled out a comprehensive battery of psychological questionnaires at home using the software REDCap: *electronic data capture tools* (Harris et a., 2009) hosted at CIMH between day 1 and day 8 consisting of more than 30 questionnaires. Relevant questionnaires for this field test were: Revised Becks-Depression Inventory (BDI-II; Kühner et al., 2007), Chronic pain grade (Korff et al., 1992), Widespread-pain-Index and Symptom Severity Scale (Frederick Wolfe et al., 2010).

In the following section some of the procedures are explained in more detail since they build the basis of the grading system operationalization of this study.

### 2.3 Data acquisition

#### 2.3.1 Overview of clinical measures for grading system application

On the first study day, we used a structured psychological interview (Mini-DIPS) to assess psychological disorders. Further, we recorded all coexisting medical conditions, including nociceptive and neuropathic pains as well as psychological disorders and sleep quality. Evoked pain hypersensitivity was examined by five outcomes of the standardized DFNS QST protocol (Rolke, Baron, et al., 2006) described in detail below, namely: Cold- and heat pain threshold, mechanical pain threshold to pinprick, pressure pain threshold and dynamic mechanical allodynia. PainDETECT questionnaire was used for checking a ‘history of hypersensitivity’. A ‘history of comorbidities’ (fatigue, cognitive problems and sleeping disturbances) was examined based on the symptom severity scale from the American College of Rheumatology (Wolfe et al., 2010). Depression was also assessed with the revised Beck depression inventory (BDI-II; Kühner et al. 2007). We used the 7-item chronic pain grading questionnaire (CPG-Q, von Korff et al., 1992) to score pain intensity (3 items) and pain interference (4 items).

#### 2.2.2 Quantitative Sensory Testing

The QST session took place at the second on-site visit, after venipuncture, an evaluation of the 7-days EMA recording and a neurofeedback session. QST was applied to the dorsum of the dominant hand of all FMS subjects (a remote, not necessarily painful area), because a previous QST study had identified signs of hyperalgesia in that location as indicator of widespread hypersensitivity (Blumenstiel et al. 2011, Gerhardt et al. 2016). In CRPS subjects, QST was performed on the affected limb: dorsum of the hand n=9, dorsolateral foot n=2. In OA and PNI patients, QST was done in the painful area.Due to time limitations we slightly changed the original DFNS QST protocol (Rolke, Magerl, et al., 2006). In short, we tested (C- and A-delta fiber mediated) cold and warm detection as well as pain thresholds with the “thermal sensory analyzer” of Medoc (Ramat Yishai, Israel), with a thermode size of 9cm^2^. Baseline temperature for each test was 32°C with a cooling or heating ramp rate of 1°/second, respectively. Cutoff temperatures were 0°C for cooling and 50°C for heating to avoid skin damage. Cold and heat pain thresholds (CPT and HPT) were determined by asking the subjects to press a stop button as soon as they perceived any nociceptive component (eg, stinging, aching, burning) in addition to cooling or warming, respectively. After they terminated each stimulus, temperature returned to baseline. The final CPT and HPT raw score is the mean of three threshold determinations, respectively.

Mechanical detection threshold (MDT), was tested with a standardized set of optic fiber based von Frey filaments (OptiHair_2_-Set, MRC Systems, Germany) that exert forces upon bending between 0.25 and 512mN graded by a factor of 2 (1–2s contact time). The contact area of the von Frey filaments with the skin was of uniform size and shape (rounded tip, 0.5mm in diameter) to avoid sharp edges that may facilitate nociceptor activation. Corresponding to the “method of limits”, five threshold determinations were made, each with a series of descending and ascending stimulus intensities (starting at a force of 16 mN). The final MDT was the geometric mean of these five series and displays the functions of A-beta fibers.

Next, we measured mechanical pain threshold (MPT) to determine A-delta mediated hyper- and hypoalgesia to pinprick stimuli. For the measurement we used a set of calibrated weighted pinprick stimulators with cylindrical tip of ϕ 0.25mm applying the forces of 8–512mN in a factor of 2 progression (“The Pinprick”; MRC Systems, Heidelberg, Germany). Subjects were asked to indicate whether they perceived a sharp sensation. Based on this decision, 5 just sub- and 5 just suprathreshold stimulus intensities were determined. The threshold was then the geometrical mean value of those stimulus intensities.

For testing A-delta mechanical pain sensitivity (MPS), only three (8mN, 64 mN and 512 mN) instead of seven pinprick stimulator intensities (MRC Systems, Heidelberg, Germany) used in the DFNS protocol were applied for three times each in a randomized order. Subjects were instructed to rate the sensation of each stimulus on a NRS ranging from 0 (no pain) to 100 (most intense pain imaginable). The subjects were asked to use a value higher than zero if they experienced any (even slight) sharp sensation. Dynamic mechanical allodynia (DMA) was tested by applying three different non-noxious stimuli to the testing area: Brush, Q-tip, and cotton wool tip were moved over the skin for 1–2cm. This procedure examines A-beta fiber mediated pain sensitivity to stroking light touch. Subjects again rated their pain on the NRS ranging from 0-100 and were instructed that any kind of sensation describing activation of nociceptors (‘sharp’, ‘burning’, ‘aching’) was defined as “pain” and was supposed to be rated with a value higher than zero. A small constant of 0.1 was added to each rating before calculating the geometrical mean value for the ratings for pinprick stimuli (mechanical pain sensitivity) and for non-noxious stimuli (DMA).

To assess temporal pain summation the Wind-up ratio (WUR) was tested: Numerical ratings within five alternating trains of a single pinprick stimulus (a) and a series (b) of 10 repetitive pinprick stimuli (at a rate of 1 Hz in an area of 1cm) were applied to calculate WUR as the ratio: mean ratings (b)/mean ratings (a). Wind-up is a measure of a frequency dependent increase in excitability of spinal cord neurons that reaches a plateau after about five stimuli (Herrero et al., 2000).

The last step of the SUPER QST protocol was the assessment of the pressure pain threshold (PPT) with a pressure algometer (FPN 100 [0–10kg scale]; Wagner Instruments, Riverside, CT, USA) with a round tip (probe diameter of 1.1cm, exerting forces up to 10kg/cm2 corresponding to ∼1000kPa). A continuous ascending ramp rate of 50 kPa/s (∼0,5 kg/ cm²) was applied manually until subjects indicated the first nociceptive sensation. The arithmetic mean of 3 successive threshold determinations was calculated as PPT. PPT is the only QST measure for deep pain sensitivity, most probably mediated by muscle C- and A-delta fibers (Rolke, Magerl, et al., 2006).

Due to time restrictions the thermal sensory limen procedure and the vibration threshold were removed from the QST protocol of SUPER. All other QST measures follow the procedure of the original protocol (Rolke, Baron, et al., 2006). Calculation of *Z*-values, thus referencing QST values to the existing reference data corresponding to age, gender, and test site, was done using Excel-based automated analyses of QST parameters (analogue to “Equista” by Casquar GmbH, Bochum, Germany).

#### 2.2.3 Pain drawings

Pain drawings were acquired on an Android device (Android Galaxy Tab S6) with a custom stylus pen and the “Squid” app (Version: 3.5.0) at the first on-site visit. Participants were asked by the neurophysiological examiner to mark all painful areas of their body on a provided body figure (two schematics - anterior and posterior view). Both the front and the back of the body had an own figure. The created pain drawings were extracted as image files (PNG format).

##### Pain area extraction

ImageJ (release: 1.53 k, Java 1.8.0_172) was used for the processing and analysis of pain drawings. Images for the front and rear-view were analyzed as follows. All participants reported the location of their most relevant pain in prior. For the sake of comparability, all pain drawings were analyzed with the most relevant pain located on the right side. For this, drawings where the main pain area was reported on the left side of the body were flipped horizontally. All image files were converted to binary images, while ensuring that all drawn areas are included. The outline of the body figure on the pain drawings was subtracted by the original body figure template image and all drawn area outside the body outline were filtered out. To remove any leftover pixel artefacts, a sequence of 3 binary erosion processes (removing the most outer pixels on the edges of each area) and a 2×2 pixel median filter were applied. To account for the reduced area due to erosion, 3 consecutive dilate processes (adding pixels to the edges of an area) were performed.

##### Group area sum

For the group sum of pain areas, the previously processed images were added to a stack. A Z-axis projection was performed by adding image slices from the stack yielding a 32-bit image of overlapping pain drawings. This image was converted to 8-bit (intensity: 0 – 255), which turns the brightest intensity of the acquired 32-bit image to the highest value intensity value of an 8-bit image (255). To ensure that the same ratio of intensities is preserved across study groups with different sample sizes, the 8-bit image was divided by the calculated ratio between the maximal theoretical intensity of a 32-bit image (255* number of slices i.e. participants) and the maximum observed intensity.

For presentation, the template was added to the group sum and a 2×2 gaussian blur filter was applied for smoothing. The image’s Look-Up Table was converted to “Fire”.

##### Pain area analysis

The area of the extracted pain areas was calculated by counting the number of pixels and relating it to the area of the filled-out body template. The areas were calculated with the “Analyze” function in ImageJ.

#### 2.4 Implementation of the grading system for ‘nociplastic pain’

We implemented data analysis according to the decision tree as follows:

Step 1 (pain chronicity) was scored with a cutoff of 3 months as in ICD-11 (Treede et al., 2015). In Step 2 we qualitatively analyzed pain drawings and the Widespread Pain Index (Wolfe et al., 2010) to assess pain spread (widespread/regional pain distribution). The 2010 ACR criteria for FMS provide a cutoff for widespread pain (WPI≥ 7). The 2019 IASP classification (M. Nicholas et al., 2019) suggests to define chronic widespread pain as diffuse musculoskeletal pain in at least 4 of 5 body regions and in at least 3 or more body quadrants (as defined by upper–lower/left–right side of the body) and axial skeleton (neck, back, chest, and abdomen). There is no published criterion for regional vs. discrete pain, although this term is used in the Budapest criteria for CRPS (Harden et al., 2010). For OA, pain was classified as regional, when there was no affected joint in one of the painful regions. For PNI, pain was classified as regional when it exceeded known anatomical distributions of innervation territories of the affected nerves. Step 3a and 3b call for examination if nociceptive or neuropathic pain mechanisms fully explain the patient’s pain. Nociceptive pain was graded as accounting fully for the pain if pain was restricted to the affected tissue (mostly joints). Neuropathic pain was graded as accounting fully for the pain if pain was restricted to the innervation territory of the affected nerve or spinal segment. Step 4 (clinical evidence for evoked pain hypersensitivity) was graded according to four QST outcomes (see above). Cold- and heat-pain thresholds, mechanical-(pinprick) and (blunt) pressure pain thresholds were used as operationalization of hypersensitivity when they exceeded a Z-score of 1.96 compared to reference data of the German research network of neuropathic pain (DFNS; Rolke, Magerl, et al., 2006). Dynamic mechanical allodynia was included as another sign of pain hypersensitivity. If at least one of the 5 phenomena was present, the participant was classified as having evoked pain hypersensitivity.

Step 5a (history of hypersensitivity) was scored according to the three items of the PainDETECT questionnaire (PD-Q) (Freynhagen et al., 2016): (a) Is light touching (clothing, a blanket) painful in this area? (b) Is cold or heat (bath water) in this area occasionally painful? (c) Does slight pressure in this area, e. g. with a finger, trigger pain? The scale of the PD-Q ranges from 0= ‘never’ to 5 = ‘very strongly’. In line with the proposed grading criteria we decided that a value of at least 2 = ‘slightly’ should be considered as fulfillment of the respective item. Step 5b (history of comorbidities) was based on the symptom severity scale from the American College of Rheumatology (Wolfe et al., 2010). The severity of the three symptoms is rated on a scale from 0 = ‘no problem’ to 3 = ‘severe problems. The minimum non-zero score of at least 1 = ‘mild’ was considered to indicate the respective comorbidity. If at least one comorbidity was reported, this was scored as present. Inclusion of this step in the decision tree has been criticized (Cohen, 2022), so we present results for each of the steps individually, and we add a critical discussion of the structure of the decision tree.

#### 2.5 Data Evaluation

Our main analyses focus on chronic primary vs. secondary pain; additionally, we present descriptive findings separately per disease. We provide means and SEM to illustrate precision of estimation of central tendency, plus medians and ranges to illustrate biological variability as appropriate. Since normal distribution was violated for some variables, we used non-parametric statistical tests. For statistical comparison of chronic primary pain and chronic secondary pain subjects Mann-Whitney U-test was used for continuous and Fisher’s exact test or chi²-test were used for categorical outcomes, respectively. We considered results with a p-value < 0.05 significant.

## 3. Results

### 3.1 Demographics and clinical evaluation

Demographics of these two and four groups respectively are shown in Table 1. We found a very strong female predominance in FMS and CRPS (82-90%), whereas there were 32-50% males in the OA and PNI group. There was no significant difference in age ranges, but PNI patients tended to be younger than the other three groups. The median of BMI in all 4 groups showed a pre-obesity stage. All patients suffered from chronic pain, with pain durations between 5 months and 43 years. Pain severity, as represented by the combination of pain intensity and disability (Treede et al., 2019) was higher in primary than secondary pain (*p* =.008, d = .61 and *p* =.008, d = .62, respectively), which was mostly due to low values in OA. In Widespread-pain-Index assessment primary pain patients on average reported 6 painful areas more compared to secondary pain patients (*p* <.001, d =1.06). Further, primary pain patients reported a higher psychological burden compared to secondary pain patients with significant differences in symptom severity score (*p* =.001, d = .79) due to high values in FMS. CRPS had similar values as OA and PNI. Likewise, BDI-II scores were higher in primary than secondary pain (*p* =.007, d = .63) due to lower burden in OA.

**Table 1.** Demographics and clinical characteristics.

|  | Chronic pain types (ICD-11) |  |  |  |  | Sub-diagnoses |  |  |  |
| --- | --- | --- | --- | --- | --- | --- | --- | --- | --- |
|  | CP1 | CP2 | *z* | *P* | *d* | FMS | CRPS | OA | PNI |
| *N* | 52 | 29 |  |  |  | 41 | 11 | 21 | 8 |
| *N* Female (%) | 46 (88.5%) | 18 (62.1 %) |  |  |  | 37 (90.2%) | 9 (81.8%) | 14 (66.7%) | 4 (50.0%) |
| Age (years) | 51.6 ± 1.2<br>53.5, 28-63 | 49.8 ± 2.3<br>50.0, 20-65 | -.128 | .898 | .028 | 51.9 ± 1.4<br>55.0, 28-62 | 50.6 ± 2.4<br>50.0, 34-63 | 52.6 ± 2<br>52.3, 21-65 | 42.4 ± 5.4<br>44.0, 20-62 |
| BMI | 29.6 ± 1.0<br>28.1, 19-45 | 29.0 ± 1.1<br>29.4, 19-43 | -.158 | .875 | .035 | 29.3 ± 1<br>27.7, 19-45 | 30.9 ± 2<br>28.7, 19-42 | 29.3 ± 1<br>29.3, 19-40 | 28.2 ± 2<br>25.3, 22-43 |
| Pain duration (years) | 17.6 ± 3.2<br>12.6, 0.4-43 | 13.3 ± 2.3<br>9.4, 0.6-33 | -.742 | .458 | .165 | 22.2 ± 4<br>19.9, 0.4-43 | 3.0 ± 1<br>1.5, 0.7-8 | 14.5 ± 3<br>10.0, 0.6-33 | 6.2 ± 3.6<br>3.2, 2-13 |
| CPG: Pain Intensity (%), | 62.1 ± 2.4<br>66.7 | 52.4 ± 3.0<br>50.0 | -2.637 | .008** | 0.613 | 61.3 ± 2.5<br>66.7 | 64.9 ± 6.4<br>66.7 | 50.5 ± 3.3<br>50.0 | 57.5 ± 6.5<br>51.7 |
| CPG: Pain Interference (%) | 54.9 ± 3.2<br>60.0 | 41.5 ± 3.7<br>40.0 | -2.654 | .008** | 0.617 | 55.7 ± 3.6<br>60.0 | 52.8 ± 7.0<br>56.7 | 39.8 ± 4.1<br>33.3 | 45.8 ± 8.3<br>45.0 |
| Widespread-Pain-Index<br>(painful areas, range 0-19) | 11.5 ± 0.7<br>12.0 | 6.3 ± 0.8<br>6.0 | -4.206 | <.001*** | 1.057 | 13.0 ± 0.7<br>13.0 | 6.2 ± 1.1<br>6.0 | 6.8 ± 0.9<br>7.0 | 5.0 ± 1.8<br>3.5 |
| Symptom Severity Score | 7.2 ± 0.3<br>7.0 | 5.3 ± 0.4<br>5.0 | -3.309 | .001*** | 0.791 | 7.7 ± 0.4<br>8.0 | 5.5 ± 0.7<br>6.0 | 5.0 ± 0.5<br>5.0 | 6.1 ± 0.9<br>6.5 |
| BDI-II Score | 21.0 ± 1.3<br>20.0 | 15.0 ± 2.1<br>13.0 | -2.711 | .007** | 0.632 | 21.3 ± 1.4<br>21.0 | 19.4 ± 3.1<br>17.0 | 13.1 ± 2.3<br>12.0 | 19.8 ± 4.0<br>19.0 |

Values present M ± SEM in the upper line and Median in the bottom line; for age, bmi and pain duration we additionally report the range (bottom). CP1 (chronic primary pain): fibromyalgia (FMS) or complex regional pain syndrome (CRPS), CP2 (chronic secondary pain): osteoarthritis (OA) or peripheral nerve injury (PNI). For CP1 vs. CP2 we present U-Test z-score, asymptotic significance, and effect size (Cohen); BMI = body mass index (normal: 24-30, obese: >35), CPG = Chronic pain grade (Von Korff et al., 1992). BDI-II: revised Beck depression inventory (minimal: 0-13, mild: 14-19, moderate 20-28, severe: 29 and above)

### 3.2 Regional vs discrete pain distribution

Fig. 2 shows the averaged pain drawings for all four diseases. In FMS (Fig. 2A), there was a high incidence of pain in all quadrants as well as axial pain indicating widespread pain. Note however, that any specific body location was painful in less than half of FMS patients. In CRPS (Fig. 2b), pain was reported for the entire hand and much of the forearm, indicating regional pain. There were 9 CRPS I with bone or joint injuries and two CRPS II patients with nerve injury. The upper limb was affected in most cases (bilaterally in 2 subjects and with additional lower limb affected in 2 cases). In OA (Fig. 2c), the most frequent location was bilateral knee joints (n =16), shoulder joints (n= 9) and zygoapophyseal joints (n =19) being affected. PNI (Fig. 2d) affected one of the three distal hand nerves (median, radial, ulnar) in most cases; the individual pain distributions were discrete and consistent with the injured nerve which is not visible in the average. Many patients with CRPS, OA or PNI reported pain in the lower back in addition to their main pain condition (see rear views in Fig. 2b-d). The total extent of painful body surface was significantly different between primary and secondary pain patients (Md = 21 % vs Md = 7%, p <.001), due to high values in FMS (23%) compared to 10% in CRPS and about 7% in both secondary pain conditions.

**Figure 1:**
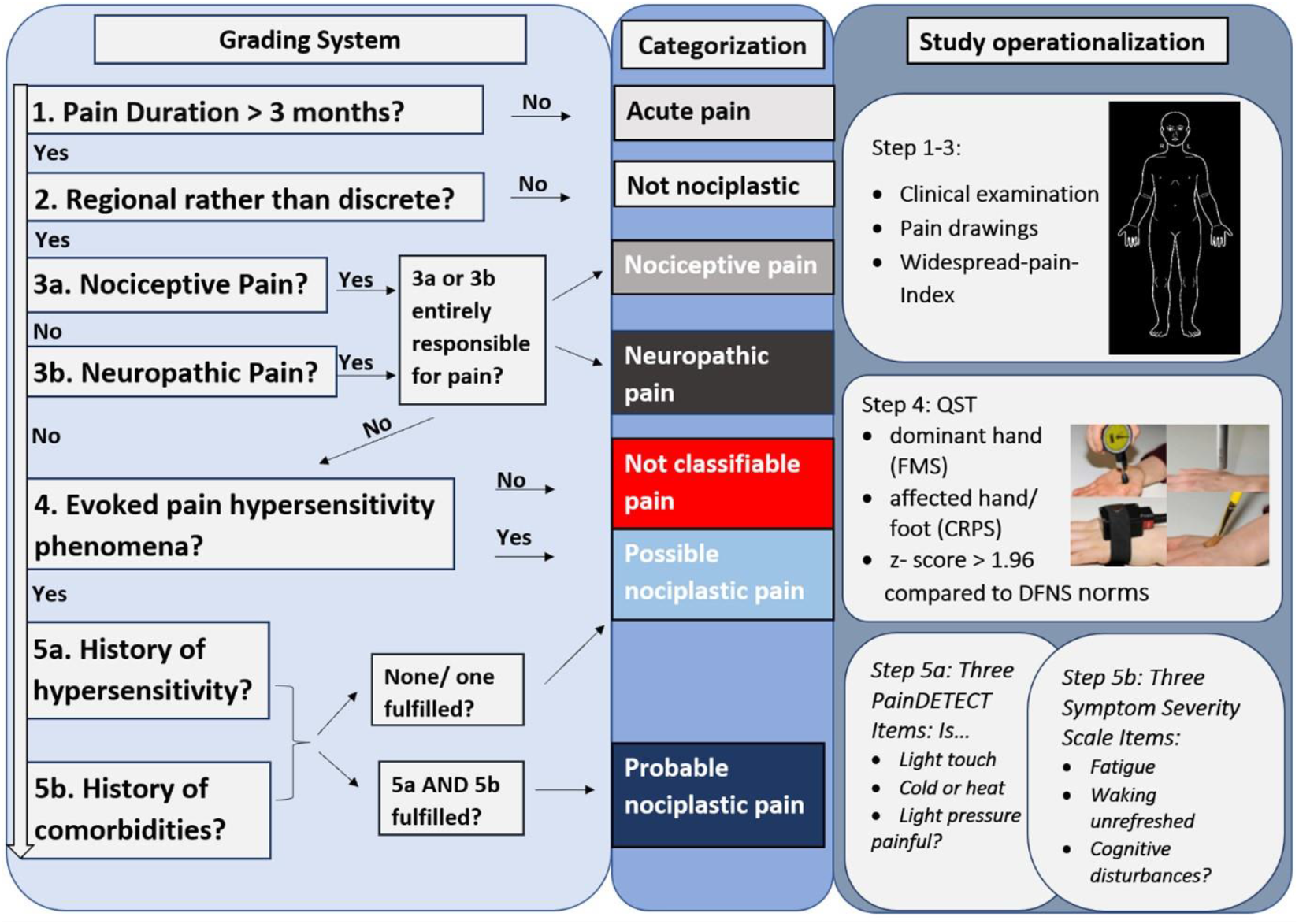
Decision tree for evaluation of evidence for nociplastic pain mechanisms. Proposed decision tree to estimate possible or probable nociplastic pain mechanisms in a given patient (Kosek et al. 2021) and its operationalization in the SUPER study.

**Figure 2.**
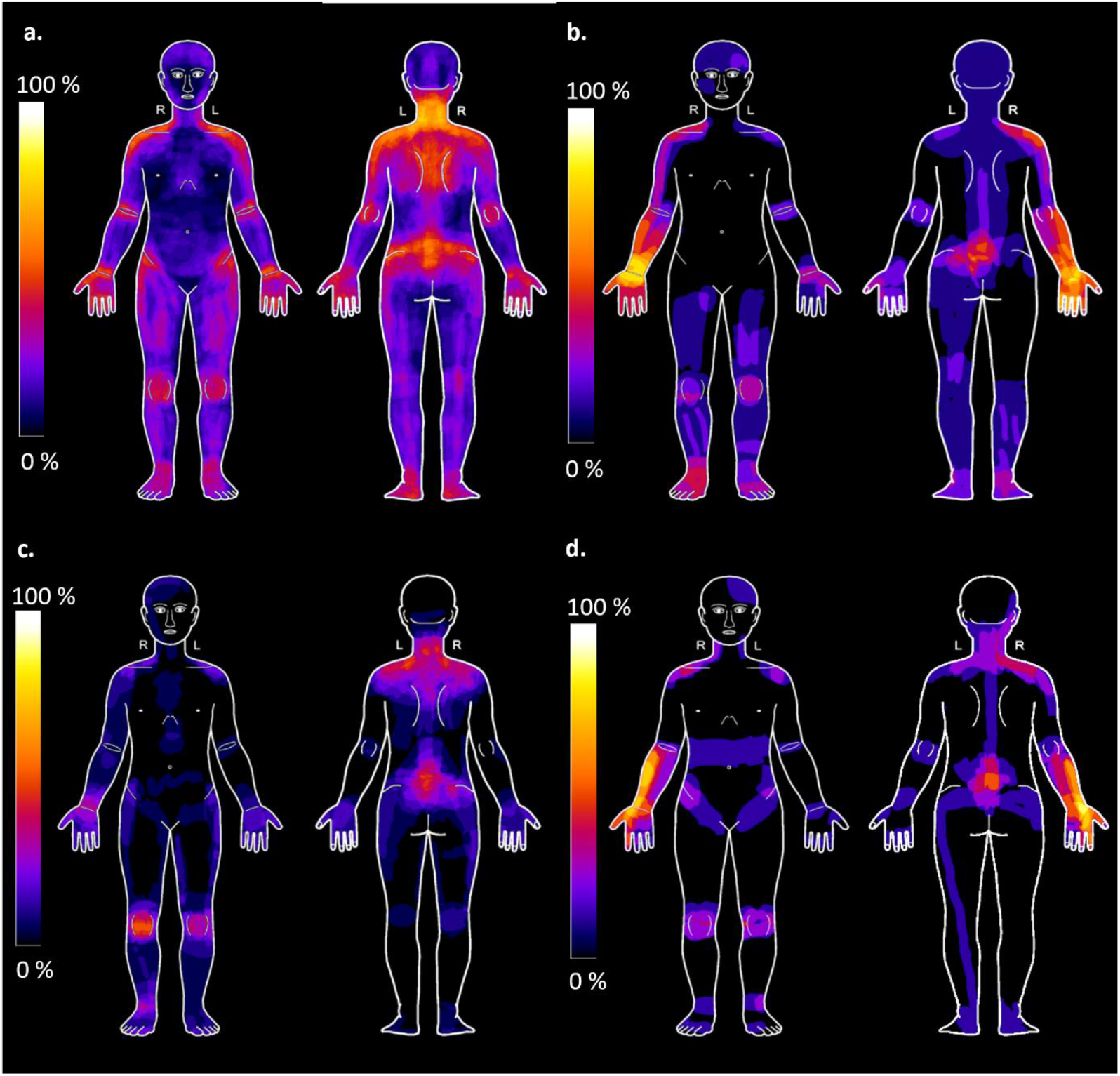
Pain Drawings for chronic primary and secondary pain conditions. Averaged pain drawings across four patient groups. If needed, the drawing was mirrored so the most painful site is on the right hand side of the body. A. Fibromyalgia syndrome (n=41, 23% of body surface), B: Complex regional pain syndrome (n=10, one missing, 10% of body surface), C: Osteoarthritis (n=20, one missing, 7% of body surface), D: Peripheral nerve injury (n=8, 7% of body surface). Heat maps indicate percentage of patients that reported pain in any given body part.

### 3.3 Coexiting Nociceptive and Neuropathic Conditions

Most patients presented with more than one pain condition. This was true for primary and secondary pain. Only 17% of patients had no comorbidities overall, with no significant difference between primary and secondary chronic pain conditions (p > 0.9). Table 2a shows co-existing painful conditions excluding the major diagnose of the respective patient group. In the FMS group, 80 % of the patients had one or more proven lesions or diseases of nervous or other tissue while 91% of the CRPS patients presented with additional (not counting the underlying trauma) pain conditions, most frequently axial pain due to spinal pathologies.

**Table 2a:**
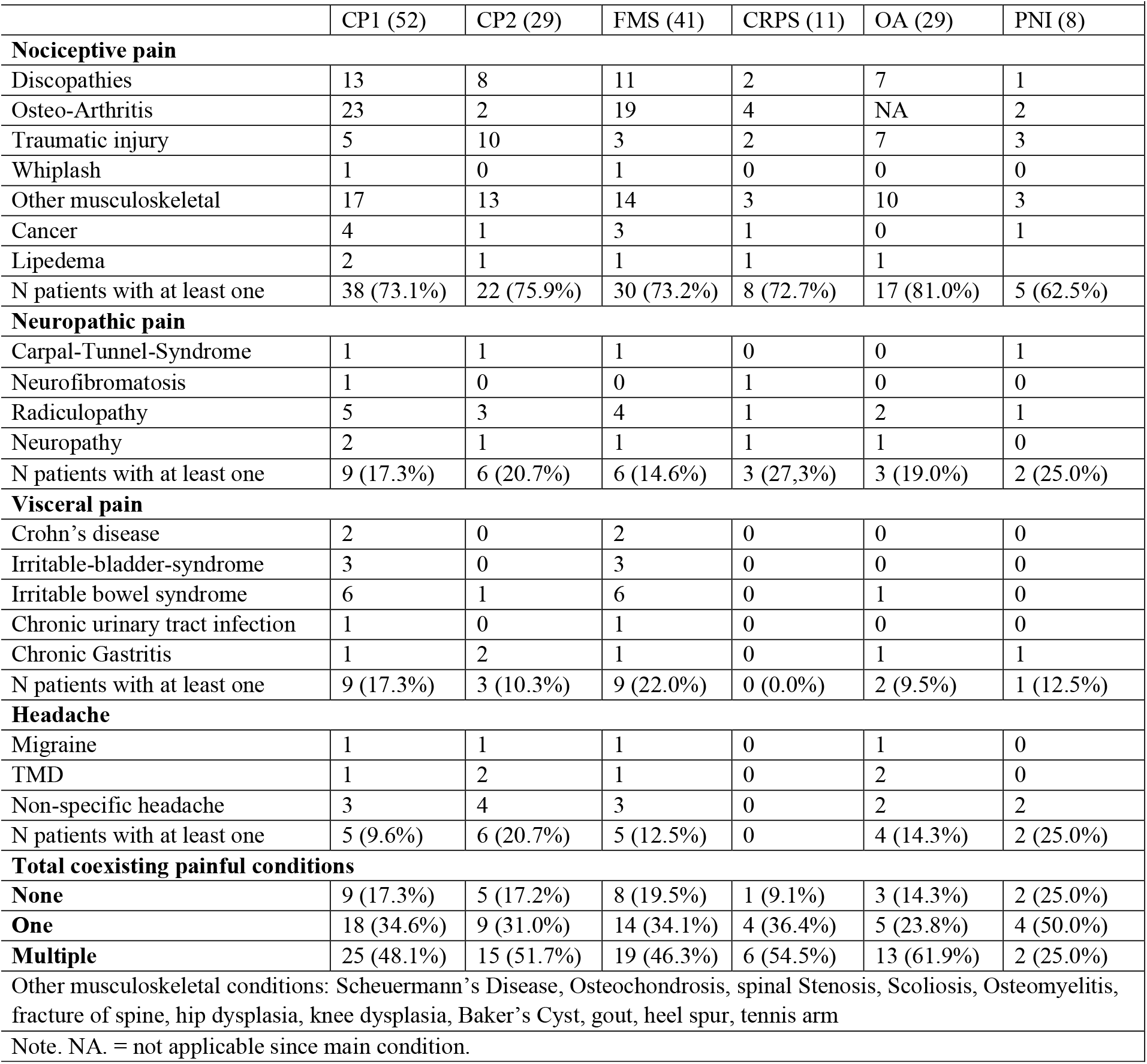
Co-existing painful conditions.

OA patients also had frequent comorbid pain (86%) as did patients with PNI (75%). Most prevalent, patients with both primary and secondary pain had additional nociceptive pain conditions and there was no difference between the groups (*p* =.99). Co-existing neuropathic pain conditions were reported in a similar frequency by 17.3% of primary pain patients and 20.7% of secondary pain patients (*p* =.94). Frequency of headaches (around 20%, lower compared to the general population (Stovner et al., 2007) did not differ significantly between chronic primary secondary pain conditions (*p* =.19). Comorbid visceral pains including primary visceral conditions were present in 22% of FMS patients resulting in 17.3% of primary pain patients affected compared to 10% in secondary pain patients. A fisher’s exact test did not show a significant difference (*p* =.52).

Table 2b presents comorbid sleep and mental disorders evaluated in an interview with a clinical psychologist. While half of the primary pain patients had a clinically relevant sleeping disorder, only about 17% of the secondary pain patients were diagnosed with one. Insomnia affected both, FMS and CRPS patients to a substantial extent (39 and 64 %, respectively). A Fisher’s exact test showed a statistically significant association between chronic pain group (CP1 vs CP2) and sleep disorder (*p* = .008). In the category of comorbid mental disorders 50% of the primary pain patients presented with a comorbid anxiety disorder and 35% with a Major Depression (MDD) or Dysthymia while in the secondary pain group 28 % suffered from a comorbid anxiety disorder and 31% from a MDD or Dysthymia. However, the comparison of overall frequency of comorbid mental disorders by a Chi² test did not show a significant relation between type of chronic pain group and mental disorders χ ² (2) =3.757, *p* =.153, ϕ = 0.22.

**Table 2b.**
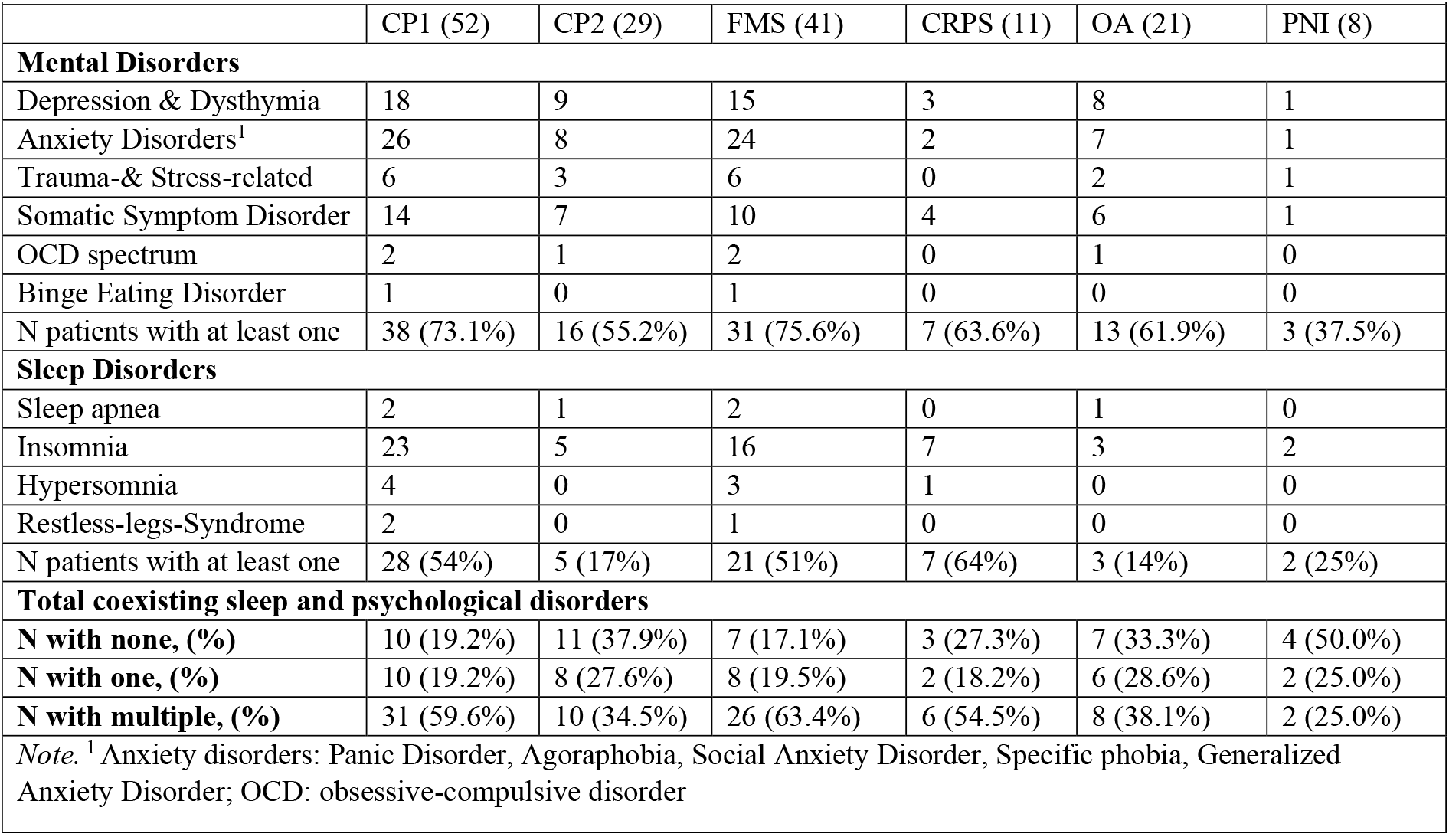
Co-existing Mental and Sleep Disorders.

| **Table 2b. Co-existing Mental and Sleep Disorders** |  |  |  |  |  |  |
| --- | --- | --- | --- | --- | --- | --- |
|  | CP1 (52) | CP2 (29) | FMS (41) | CRPS (11) | OA (21) | PNI (8) |
| **Mental Disorders** |  |  |  |  |  |  |
| Depression & Dysthymia | 18 | 9 | 15 | 3 | 8 | 1 |
| Anxiety Disorders<sup>1</sup> | 26 | 8 | 24 | 2 | 7 | 1 |
| Trauma-& Stress-related | 6 | 3 | 6 | 0 | 2 | 1 |
| Somatic Symptom Disorder | 14 | 7 | 10 | 4 | 6 | 1 |
| OCD spectrum | 2 | 1 | 2 | 0 | 1 | 0 |
| Binge Eating Disorder | 1 | 0 | 1 | 0 | 0 | 0 |
| N patients with at least one | 38 (73.1%) | 16 (55.2%) | 31 (75.6%) | 7 (63.6%) | 13 (61.9%) | 3 (37.5%) |
| **Sleep Disorders** |  |  |  |  |  |  |
| Sleep apnea | 2 | 1 | 2 | 0 | 1 | 0 |
| Insomnia | 23 | 5 | 16 | 7 | 3 | 2 |
| Hypersomnia | 4 | 0 | 3 | 1 | 0 | 0 |
| Restless-legs-Syndrome | 2 | 0 | 1 | 0 | 0 | 0 |
| N patients with at least one | 28 (54%) | 5 (17%) | 21 (51%) | 7 (64%) | 3 (14%) | 2 (25%) |
| **Total coexisting sleep and psychological disorders** |  |  |  |  |  |  |
| **N with none, (%)** | 10 (19.2%) | 11 (37.9%) | 7 (17.1%) | 3 (27.3%) | 7 (33.3%) | 4 (50.0%) |
| **N with one, (%)** | 10 (19.2%) | 8 (27.6%) | 8 (19.5%) | 2 (18.2%) | 6 (28.6%) | 2 (25.0%) |
| **N with multiple, (%)** | 31 (59.6%) | 10 (34.5%) | 26 (63.4%) | 6 (54.5%) | 8 (38.1%) | 2 (25.0%) |
| *Note.* <sup>1</sup> Anxiety disorders: Panic Disorder, Agoraphobia, Social Anxiety Disorder, Specific phobia, Generalized Anxiety Disorder; OCD: obsessive-compulsive disorder |  |  |  |  |  |  |

### 3.4 Clinically evoked hypersensitivity phenomena

We used the standardized and validated QST protocol of DFNS to assess signs of hypersensitivity. We analyzed pain thresholds for noxious cold, heat, pinprick, and blunt pressure. After normalization to sex, age and test site, standard z-scores for thermal pain were higher in FMS than CRPS, but scores for mechanical pain were higher for CRPS. OA patients had sensory gain across all four tests, while PNI had sensory loss for thermal and gain for mechanical noxious stimuli.

On an individual patient basis, 15 FMS patients showed evidence for thermal hyperalgesia (34%), and in OA this was also frequent (6/23=26%), while thermal hyperalgesia was rare in CRPS (18%) and PNI (13%). The most frequent nociceptive sensory gain was MPT in CRPS (6/11 = 55%). DMA was reported most frequently by CRPS and PNI patients (54% and 75%, respectively, Tab. 3). Thus, QST provided clinical evidence for hypersensitivity in many patients, 69% of chronic primary pain (27/41 FMS, 9/11 CRPS) and equally 69 % of chronic secondary pain (13/21 OA, 7/8 PNI). We did not find a significant relation between evoked hypersensitivity and chronic pain group in the full sample (χ ² (1) < 0.001, *p* >.9, *V* < 0.1).

**Table 3.** Evoked hypersensitivity phenomena in full sample - Signs of sensory gain in QST.

|  |  |  |  |  |  |  |
| --- | --- | --- | --- | --- | --- | --- |
|  | 1°CP (52) | 2°CP (29) | FMS (41) | CRPS (11) | OA (21) | PNI (8) |
| **Cold pain threshold** |  |  |  |  |  |  |
| None | 28 | 23 | 20 | 8 | 15 | 8 |
| One SD over DFNS norm | 13 | 5 | 11 | 2 | 5 | 0 |
| Two SDs over DFNS norm | 11 | 1 | 10 | 1 | 1 | 0 |
| **Heat pain threshold** |  |  |  |  |  |  |
| None | 27 | 16 | 20 | 7 | 10 | 6 |
| One SD over DFNS norm | 8 | 7 | 7 | 1 | 5 | 2 |
| Two SDs over DFNS norm | 17 | 6 | 14 | 3 | 6 | 0 |
| **Pressure pain threshold** |  |  |  |  |  |  |
| None | 17 | 15 | 15 | 2 | 10 | 5 |
| One SD over DFNS norm | 12 | 9 | 9 | 3 | 7 | 2 |
| Two SDs over DFNS norm | 23 | 5 | 17 | 6 | 4 | 1 |
| **Mechanical pain threshold (pinprick)** |  |  |  |  |  |  |
| None | 19 | 16 | 15 | 4 | 11 | 5 |
| One SD over DFNS norm | 16 | 9 | 15 | 1 | 7 | 2 |
| Two SDs over DFNS norm | 17 | 4 | 11 | 6 | 3 | 1 |
| **Dynamic mechanical allodynia** |  |  |  |  |  |  |
| No | 44 | 21 | 39 | 5 | 19 | 2 |
| Yes | 8 | 8 | 2 | 6 | 2 | 6 |
| **Percentage fulfillment of overall criterion “subjectively reported history of hypersensitivity”<sup>1</sup>** |  |  |  |  |  |  |
| N fulfilled | 36 (69.2%) | 20 (69.0%) | 27 (65.9%) | 9 (81.8%) | 13 (61.9%) | 7 (87.5%) |
| *Note.* <sup>1</sup> Criterion was fulfilled if patient showed at least one out of five pathological hypersensitivity signs. |  |  |  |  |  |  |

### 3.5 History of hypersensitivity and comorbidities

The criterion “history of hypersensitivity” was assessed with items of the painDETECT questionnaire. According to our operationalization we assumed the criterion to be fulfilled if a subject rated at least one of three painDETECT items ‘slightly’ or above. Table 4a shows the rating of all subjects investigated. Considering the total sample, about 83% of subjects in both groups, CP1 and CP2 would fulfill the criterion. The criterion “history of comorbidities” was assessed with items of the symptom severity scale. According to our operationalization we assumed the criterion to be fulfilled if a subject indicated to have ‘mild’ to ‘severe’ problems with at least one of three potential comorbidities. Table 4b shows, that this criterion was fulfilled by all subjects investigated.

**Table 4a.**
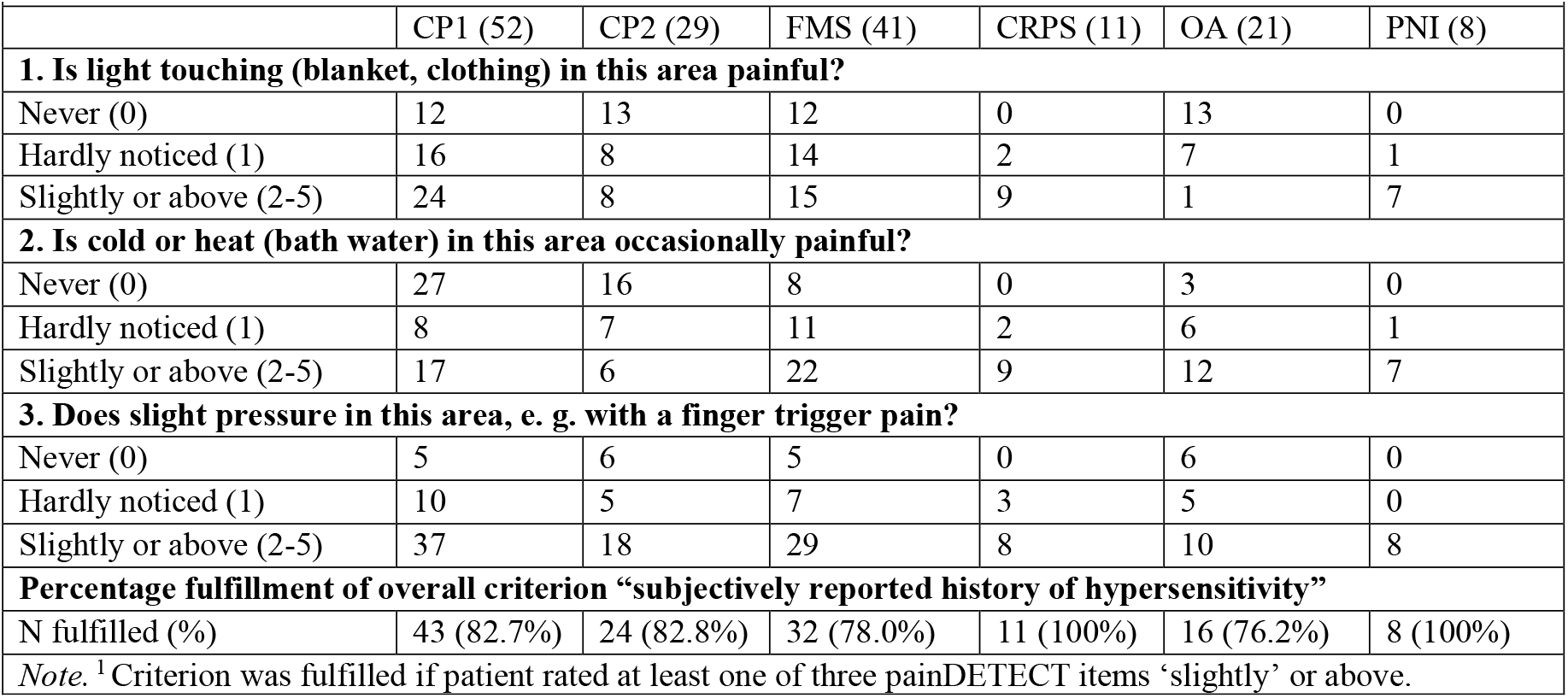
History of hypersensitivity phenomena – Items of painDETECT Questionnaire.

**Table 4b.**
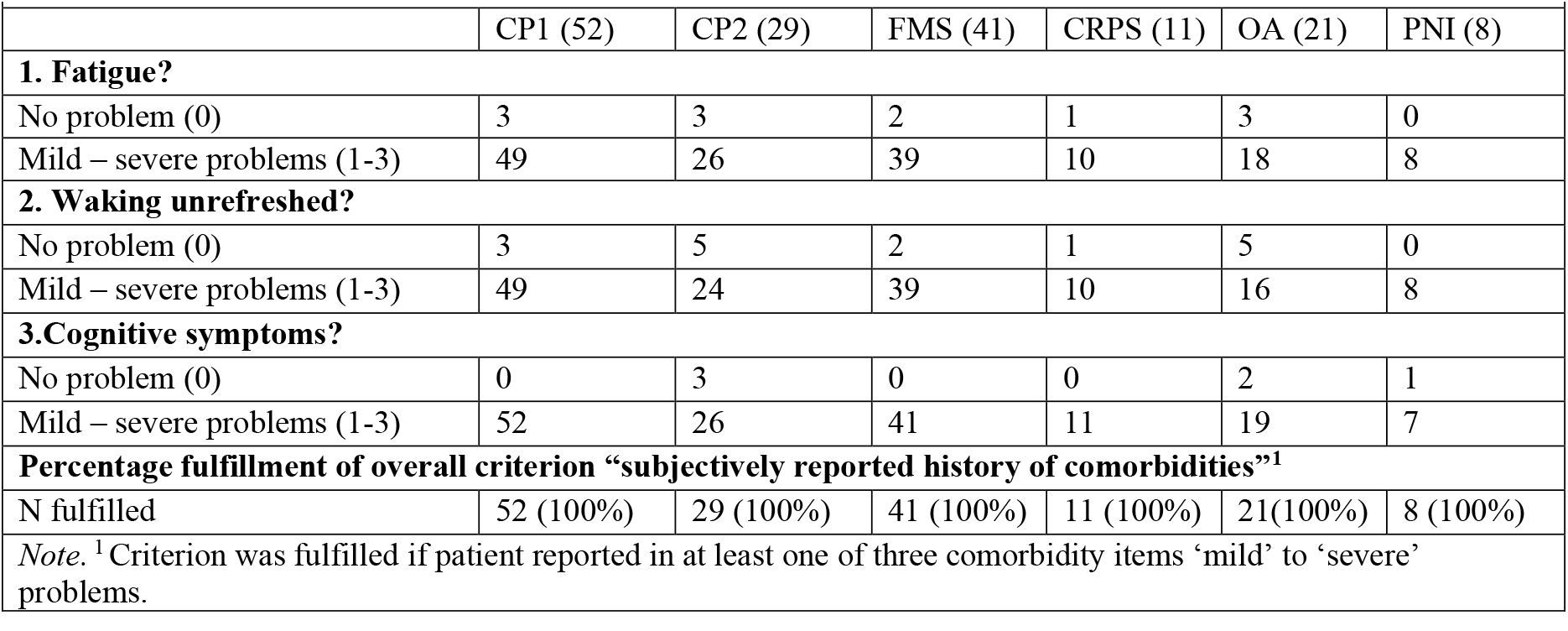
History of comorbidities – Items of Symptom Severity Scale.

### 3.6 Classification according to the nociplastic grading system

Based on these data, we classified all patients according to the nociplastic pain decision tree (Tab. 5, Fig. 5). All patients fulfilled the duration criterion. In pain spread analyses 38% of the OA and 50% of the PNI patients showed discrete pain and therefore dropped out of further steps in the grading system. 36 of the FMS and 12 participants in the OA group had widespread pain. In a minority of the FMS group the pain extent was regional (15%) as well as in one OA subject. Thus, in all chronic primary pain patients but only in about half of the chronic secondary pain patients it was evaluated if another nociceptive or neuropathic mechanism could fully explain the pain. The pain of 12 OA patients could be entirely explained by nociceptive mechanisms due to their arthritis condition. Furthermore, the pain of 75% of the remaining PNI subjects could be explained by their respective nerve injury. This left all primary pain subjects and one subject in each of the two secondary pain groups for the evaluation of clinical hypersensitivity. In FMS, 14 subjects (34%) did not show a pathological hypersensitivity according to DFNS norms in QST, while in CRPS this was the case in 2 subjects (18%). In the proposed grading system these primary pain patients therefore end up classified with ‘not classifiable pain’ (Fig. 4). This label was significantly associated with the chronic primary pain group (Fisher’s exact *p* <.001). Both secondary pain patients fulfilled the evoked hypersensitivity criterion. In step 5a, 5 FMS (12%) subjects did not report a history of hypersensitivity in the painDETECT items, while all CRPS, OA and PNI participants did. A Fisher’s exact test did not a significant relation between the label ‘possible nociplastic pain’ and chronic pain group (*p* = .154). In line with the high rates of psychological disorders, all subjects in both groups reported to suffer from at least one psychological comorbidity (Tab 4b). In consequence based on our operationalization about 60% of the CP1 and only 7% of the CP2 patients were classified to have ‘probable nociplastic pain’ (Tab. 5). In line with our hypotheses, a Fisher’s exact test revealed a significant association between chronic pain group (CP1 vs CP2) and the classification of a “probable nociplastic pain mechanism” (*p* < .001). Still, in the primary pain group 82% of the CRPS but only 54% of the FMS subjects were classified to have ‘probable’ nociplastic pain (Fig. 4), however this difference in sub-diagnoses in the CP1 group was not significant (Fisher’s exact *p* = .165).

**Figure 3:**
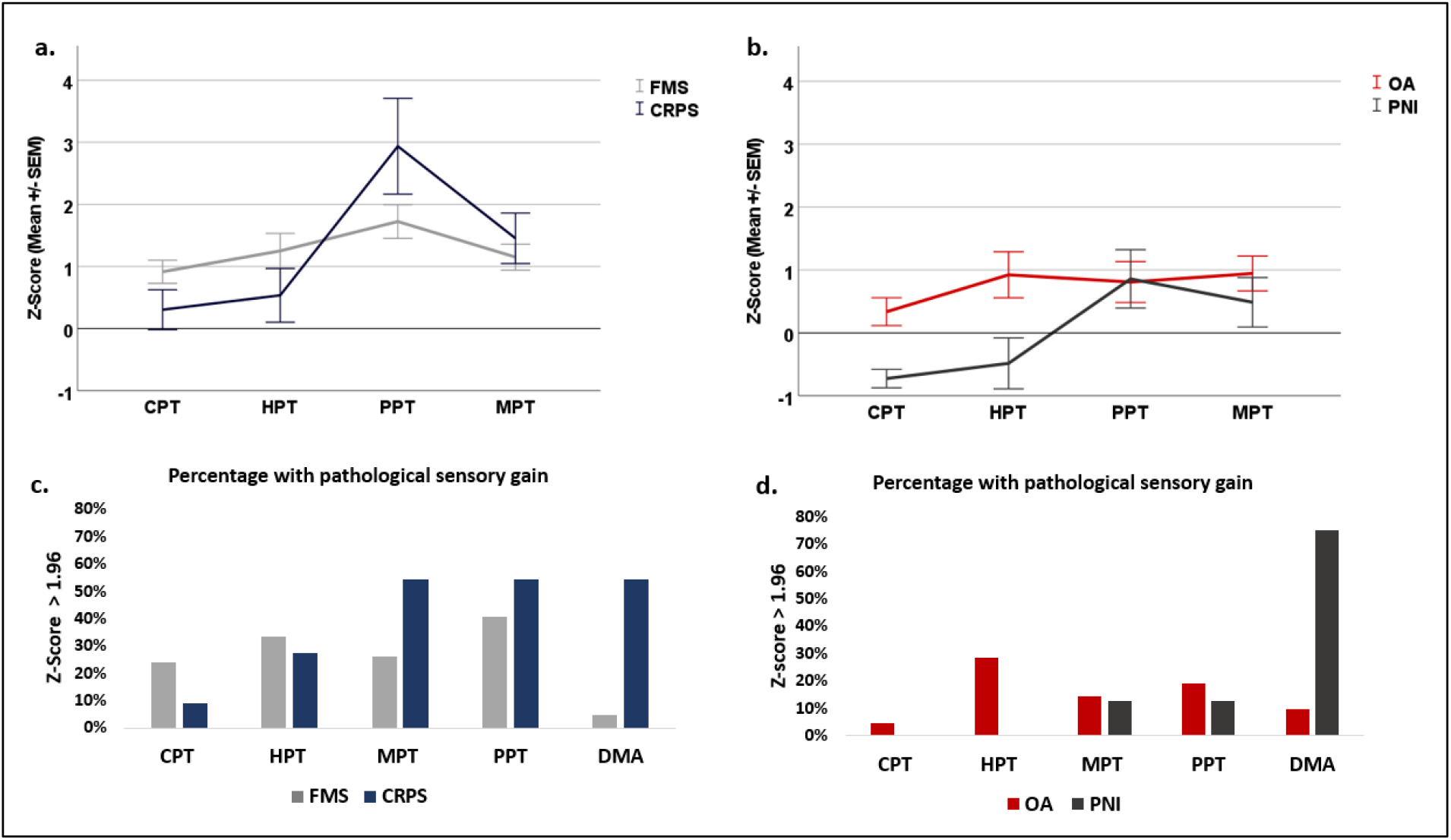
Signs of evoked pain hypersensitivity for chronic primary and secondary pain conditions. Pain thresholds were determined according to the QST protocol of DFNS. Top row: z-scores normalized to DFNS reference data (mean ± SEM). Bottom row: percentage of patients with significant nociceptive sensory gain (z>1.96). A: chronic primary pain conditions (FM: fibromyalgia syndrome (n = 41), CRPS: complex regional pain syndrome (n = 11). B: chronic secondary pain conditions (OA: osteoarthritis (n = 21), PNI: peripheral nerve injury (n = 8). CPT: cold pain threshold, HPT: heat pain threshold, PPT: pressure pain threshold, MPT: mechanical (pinprick) pain threshold, DMA: dynamic mechanical allodynia. Note that some neuropathic pain patients had significant small fiber sensory loss that is not evaluated here.

**Figure 4:**
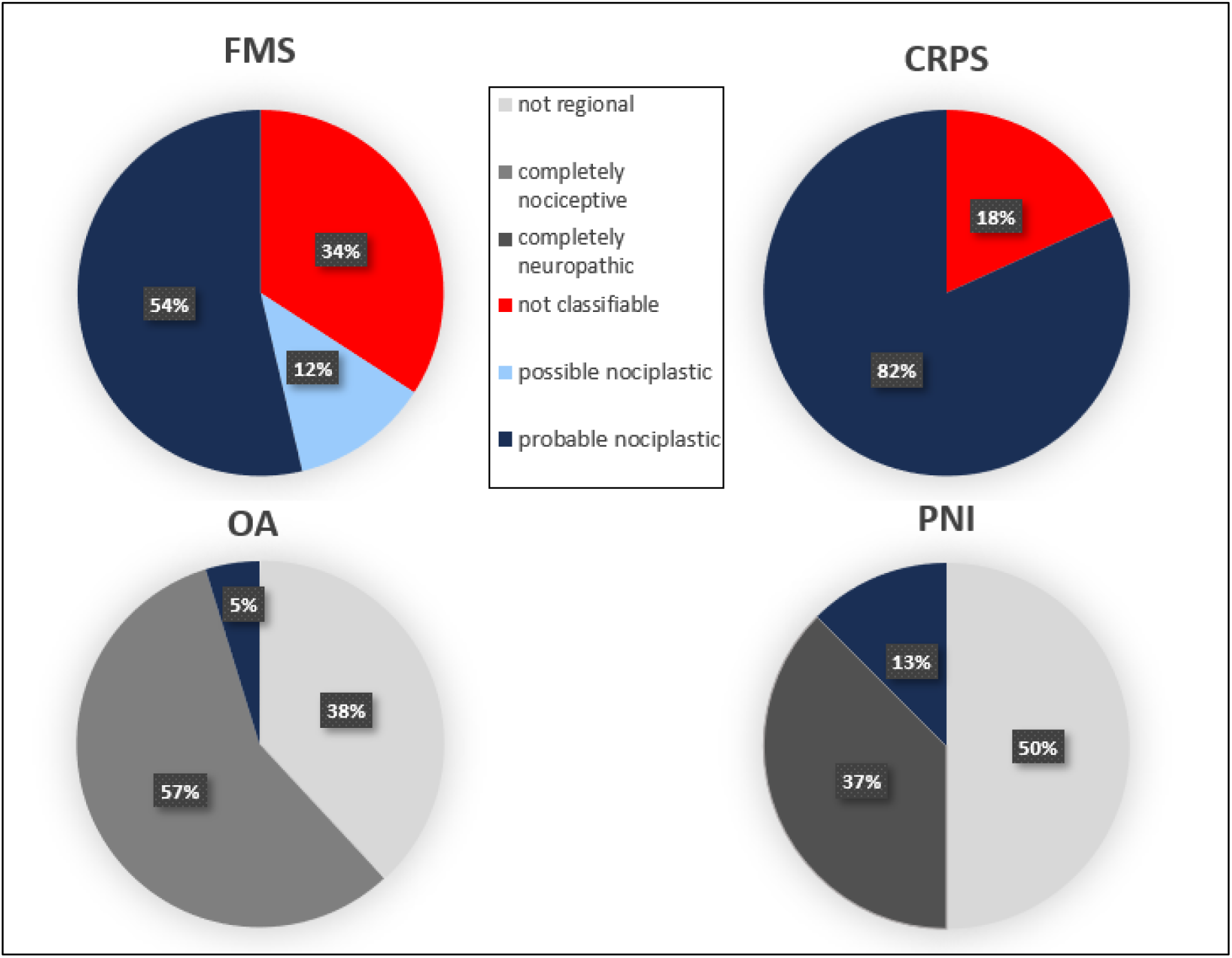
Outcome of nociplastic pain grading for four patient populations. Pie charts depict percentage of patients that are classified as ‘not regional’, completely nociceptive or neuropathic (grey shades), not classifiable pain (red), possible nociplastic pain (light blue) and probable nociplastic pain (dark blue) according to the published grading system (Kosek et al. 2021). FMS: Fibromyalgia syndrome (n=41), CRPS: Complex regional pain syndrome (n=11), OA: Osteoarthritis (n=21), PNI: Peripheral nerve injury (n=8).

**Table 5.** Application of the grading system for nociplastic pain.

| **Table 5. Application of the grading system for nociplastic pain** |  |  |  |  |  |  |
| --- | --- | --- | --- | --- | --- | --- |
|  | CP1 (52) | CP2 (29) | FM (41) | CRPS | OA (21) | PNI (8) |
| **Step 1\| Pain duration > 3 months** |  |  |  |  |  |  |
| No | 0 | 0 | 0 | 0 | 0 | 0 |
| Yes | 52 | 29 | 41 | 11 | 21 | 8 |
| To next step? | 52 (100%) | 29 (100%) | 41 (100%) | 11 | 21 (100%) | 8 (100%) |
| **Step 2\| Spatial extent of major pain condition** |  |  |  |  |  |  |
| Discrete | 0 | 12 | 0 | 0 | 8 | 4 |
| Regional | 16 | 5 | 5 | 11 | 1 | 4 |
| Widespread | 36 | 12 | 36 | 0 | 12 | 0 |
| To next step? | 52 (100%) | 17 (58.6%) | 41 (100%) | 11 | 13 (62) | 4 (50%) |
| **Step 3\| Other pain mechanism in painful area?** |  |  |  |  |  |  |
| **a. Nociceptive?** |  |  |  |  |  |  |
| None | 18 | 2 | 11 | 7 | 0 | 2 |
| Some | 34 | 3 | 30 | 4 | 1 | 2 |
| Complete origin | 0 | 12 | 0 | 0 | 12 | 0 |
| To next step? | 52 (100%) | 5 (17.2%) | 41 (100%) | 11 | 1 (5%) | 4 (50%) |
| **b. Neuropathic?** |  |  |  |  |  |  |
| None | 45 | 1 | 36 | 9 | 1 | 0 |
| Some | 7 | 1 | 5 | 2 | 0 | 1 |
| Complete origin | 0 | 3 | 0 | 0 | 0 | 3 |
| To next step? | 52 (100%) | 2 (6.9%) | 41 (100%) | 11 | 1 (5%) | 1 (13%) |
| **Step 4\| Clinically evoked hypersensitivity?** |  |  |  |  |  |  |
| **No / Not classifiable pain** | 16 | 0 | 14 | 2 | 0 | 0 |
| **Yes / Possible nociplastic** | 36 | 2 | 27 | 9 | 1 | 1 |
| To next step? | 36 (69.2%) | 2 (6.9%) | 27 (66%) | 9 (82%) | 1 (5%) | 1 (13%) |
| **Step 5\| History of ...** |  |  |  |  |  |  |
| **a. hypersensitivity?** |  |  |  |  |  |  |
| No | 5 | 0 | 5 | 0 | 0 | 0 |
| Yes | 31 | 2 | 22 | 9 | 1 | 1 |
| To next step? | 31 (59.6%) | 2 (6.9%) | 22 (54%) | 9 (82%) | 1 (5%) | 1 (13%) |
| **b. comorbidities?** |  |  |  |  |  |  |
| No | 0 | 0 | 0 | 0 | 0 | 0 |
| **Yes/ Probable nociplastic** | 31 (59.6%) | 2 (6.9%) | 22 (54%) | 9 (82%) | 1 (5%) | 1 (13%) |

## 4 Discussion

In our field test of the grading system for musculoskeletal nociplastic pain (Kosek et al., 2021) we found a significant difference in frequency of chronic primary and chronic secondary subjects to be classified as having “possible” or “probable” nociplastic pain. Only one subject of each secondary pain group (OA and PNI) stayed in the decision tree until the examination of clinically evoked hypersensitivity, while all primary pain patients did. However, in the CP1 group, almost all CRPS subjects (82%) but only 27 out of 41 FMS subjects (66%) showed evoked hypersensitivity phenomena in QST and were classified as having ‘possible nociplastic pain’. Five FMS subjects did not report a history of hypersensitivity in step 5a. All remaining subjects (22 FMS, 9 CRPS) reported to be at least mildly affected by one of the three psychological comorbidities in step 5b. Consequently, our hypothesis that all CP1 subjects would be rated as “probable” nociplastic was not verified since 31 % of the CP1 subjects were labelled as having “not classifiable pain” in criterion four of the grading system.

### 4.1 Chronic non-nociceptive, non-neuropathic pain

The duration criterion is based on the chronic pain definition in ICD-11 (Treede et al., 2019), where three months is a temporal criterion that was agreed on by the IASP Taskforce since the concept of persistence beyond normal healing time or pain lacking a warning function is difficult to verify in most conditions (Cervero & Laird, 1991; Treede et al., 2019). Still, some recent work suggests abandoning the temporally based pain classification scheme because it does not accurately reflect the underlying principles inherent in the phenomena of pain (Loeser, 2022; M. K. Nicholas, 2022), i.e. for nociplastic pain it remains a point of investigation at which point homeostatic plasticity (Cervero & Laird, 1991) turns into a dysfunctional functional and structural plasticity in the nociceptive system. Thus, the used temporal criterion of three months is in line the18 current agreement on chronic pain but remains to be subject of critical examination.

Concurrent clear nociceptive activation in the region of pain or a neuropathic condition that fully explains the symptoms of a patient rule out the diagnosis of nociplastic pain a priori (Kosek et al., 2016; Magerl, 2022), still it is broadly assumed that nociceptive, neuropathic and nociplastic mechanisms may occur in parallel and ongoing nociceptive pain constitutes a risk factor for developing nociplastic pain (Kosek et al., 2021). For the exclusion of a neuropathic mechanism Finnerup et al. (2016) proposed an updated grading system containing an overview of neuroanatomically plausible distributions of pain symptoms and sensory signs in common neuropathic conditions. Nociceptive pain is described as pain that is proportionate to the nature and extend of non-neural tissue injury or pathology (Nijs et al., 2014). The IASP defined it as “Pain that arises from actual or threatened damage to non-neural tissue and is due to the activation of nociceptors” (IASP Terminology). Further, there are clear criteria for classification of osteoarthritis (Altman, 1991). However, some non-specific imaging findings, such as degenerative disc disease, bulging discs or minor abnormalities like leg length discrepancies, are to some extend common in the general adult population and should not carelessly be taken as definite causation of a nociceptive pain, respectively (Lumley & Schubiner, 2019).

Thus, the first three mandatory negative criteria need to be evaluated very carefully by a trained medical expert to study neuroanatomical plausibility of nociceptive or neuropathic mechanisms causing the entirety of reported pain symptoms. The criteria and tools explained above may help guide this decision.

### 4.2 Examination of pain spread warrants further elaboration

Since the descending modulating system is linked to broad parts of the body, a widespread pain pattern is indicative of its dysfunction (Treede, 2022) and thus to sensitization, the central concept of nociplastic pain. Originally, widespread pain has been defined as axial pain plus upper and lower segment pain as well as left- and right-sided pain (Wolfe et al., 1990). More recently, four out of five regions are required to be painful for the classification as widespread pain (M. Nicholas et al., 2019). We did not count abdominal pain as axial pain, but this issue is controversial.

The criteria for regional pain have not been defined as rigorously yet. Although CRPS refers to regional ongoing pain, this aspect is not elaborated in the Budapest criteria (Harden et al., 2010). For CRPS II, a criterion for regional pain could be spatial extent beyond the innervation territory of the injured nerve. But for CRPS I it is less clear, whether a regional pain clearly extends beyond the territory of referred or radiating deep tissue pain.

In the proposed criteria, Kosek et al. (2021) name regional or multifocal rather than discrete pain as indicative for nociplastic pain. This criterion lacks a clearcut definition. Lumley and Schubiner (2019) describe the pattern of centralized pain to be inconsistent with any structural disorder, such as being located on one whole side of the body or the entire arm or leg. They further add shifting pain locations or spreading from one area to adjacent regions over time to the description. Pain drawings were proposed to standardize and optimize the assessment of pain distribution in a reliable way (Nijs et al., 2021). Further, if patients report diffuse pain that is hard for them to localize precisely, this can be an indicator for referred pain linked to central sensitization (Arendt-Nielsen & Graven-Nielsen, 2003). Deep somatic pain – like visceral pain – is often associated with pain referral that includes pain and hypersensitivity radiating from the affected tissue into adjacent uninjured tissues. A broad evidence base on pain radiation pattern for nociceptive-tissue pains would be helpful to discriminate these radiation patterns from neuroanatomical illogical pain patterns that are indicative of nociplastic pain.

### 4.3 Clinical examination of hyperalgesia warrants further elaboration

The first positive sign that is mandatory to classify a patient as suffering from ‘possible nociplastic pain’ is a clinical evoked hypersensitivity at least in the region of pain (Kosek et al., 2021; Nijs et al., 2014) which encompasses allodynia, hyperalgesia and painful aftersensations. These are signs which can indirectly show sensitization of the nociceptive system (IASP terminology; Nijs et al., 2021) defined by the IASP as ‘Increased responsiveness of nociceptive neurons to their normal input, and/or recruitment of a response to normally subthreshold inputs’ (IASP Terminology).

Using established, norm-score based QST as operationalization of the criterion “evoked hypersensitivity” of the grading system led to a high drop out of FMS subjects as “not classifiable” (34%). This may indicate that the definition of a pathological gain based on DFNS QST norm score cut-off could be too strict as definition of the grading criterion four, especially when testing in a remote instead of the most painful region in the FMS group. However, testing in a remote rather than the painful region is necessary for detecting secondary mechanical hyperalgesia linked to spinal sensitization and widespread hypersensitivity related to dysfunctional brainstem controls (Treede et al., 2022). Further, it has been discussed to increase discriminative ability of the grading system (Nijs et al., 2021). Nijs et al (2010) discriminate neuropathic central sensitization (CS) from non-neuropathic CS and state that clinical examination of the latter typically reveals increased sensitivity at sites segmentally unrelated to the primary source of nociception like referred pain. However, our results show that testing in only one remote region may lead to a loss in sensitivity of the classification. Thus, hypersensitivity should optimally be tested in multiple regions. A useful addition to detect hypersensitivity in multiple areas could be a high tender point count.

### 4.4. The relevance of subjective history of hypersensitivity and psychological comorbidities

A small proportion of FMS patients (12%) did not report a history of hypersensitivity in the painDETECT items. Along with the missing evoked hypersensitivity, this could be an indicator, that some FMS patient’s prominent symptom is not hyperalgesia. However, since we only used three items of painDETECT, future studies should evaluate the use of other questionnaires where also hypersensitivity to other stimuli, such as light or odors is quantified (e. g. Central-Sensitization-Inventory; Mayer et al., 2012).

The assessment of comorbidities showed that the operationalization based on the grading system of Kosek et al. (2021) does not discriminate between patient groups, since all subjects indicated to have at least mild problems with either Fatigue, unrefreshed sleep or cognitive disturbances. It should be considered to examine a broader range of potential signs of cortical sensitization to pain, such as dysfunctional interoception and hypervigilance to pain, jumping-to-conclusions, an increased arousal level, Kinesiophobia or pain catastrophizing. Future study may detect if some of these potential markers show higher discriminative value for the suggested concept of nociplastic pain than others.

### 4.5 Adjusting the sequence of self-report and clinical evaluation

We suggest that evidence of hypersensitivity derived from clinical examination is placed too early in the decision tree. In medical diagnostics, clinical examination normally follows patient reported history (Treede et al., 2008) and hence criterion four and five should be switched. In line with the neuropathic pain grading system of 2008, a careful examination of subjective history of hypersensitivity as well as the above proposed signs of cortical sensitization to pain should rather lead to the term “possible nociplastic pain” and a following clinical examination of evoked hypersensitivity phenomena (or other potential future biomarkers) may be used to reach the classification of “probable nociplastic pain”.

### 4.6 Limitations

As a limitation, our QST testing was done in a standard location (hand) often remote from the painful region in FMS patients while it was done in the painful area in the rest of participants. Therefore, it is not straightforward possible to compare frequency of evoked hypersensitivity. Still, using this deviation from the originally proposed protocol brought up the important point of discussion about the area of clinically testing evoked hypersensitivity and the suggestion to add tender point count as a measure. Another challenge of the study was the recruitment of participants during the COVID19 pandemic, resulting in broadly differing numbers of subjects in the four respective groups. To explore the application of the grading system with sufficient statistical power future research should aim to include a higher number of CRPS and PNI patients.

### 4.7 Outlook and Conclusion

The decision tree for the likelihood of involvement of nociplastic pain mechanisms proposed by Kosek et al. 2021 separated the chronic primary pain conditions CRPS from the chronic secondary pain condition PNI, although initiating events for both conditions partly overlap. However, the prototypical disease for both chronic primary pain and potential nociplastic mechanisms, FMS, reached the level of “probable nociplastic pain” in slightly more than half of the cases. We propose that detection sensitivity of the decision tree may be improved by switching self-reported hypersensitivity and psychological comorbidities (Step 5) and investigator observed evidence for hypersensitivity (step 4); this would be consistent with common medical assessment strategies that place history before clinical examination (Treede et al., 2008). Moreover, re-introduction of tender point counts (ACR 1990 criteria; Wolfe et al., 1990) could be taken as evidence for hypersensitivity in FMS. Further, we suggest that the clinical application of the grading system will profit from a clear definition of regional and multifocal pains as well as an evidence base on pain radiation patterns for nociceptive deep-tissue pains. Moreover, further potential signs of cortical sensitization should be tested in future studies. In summary, the decision tree proposed by Kosek et al. 2021 has the advantage of providing testable positive identification criteria. Our data suggest that its current version leads to both over- and underestimation of the “nociplasticity” of chronic pain. The overestimation raises the concern that the concept of nociplastic pain may be too broad to be useful clinically. This concern is supported by existing evidence that sensitization of nociceptive pathways may occur with any acute or chronic pain conditions (Treede, 2022).

## Data Availability

All data produced in the present study are available upon reasonable request to the authors

## Acknowledgements

This study was supported by the German Research Council (DFG) in collaborative research center grant SFB 1158, subprojects B04, B09 and S01. We thank Prof. Dr. Leila Harhaus and her team for the support and expertise in patient recruitment. Further we thank our study nurse Yvonne Neu as well as the Lena Skatulla, Michelle Berkemann and Niko Möller-Grell for their help in data acquisition.

